# Kinetics of antibody responses dictate COVID-19 outcome

**DOI:** 10.1101/2020.12.18.20248331

**Authors:** Carolina Lucas, Jon Klein, Maria Sundaram, Feimei Liu, Patrick Wong, Julio Silva, Tianyang Mao, Ji Eun Oh, Maria Tokuyama, Peiwen Lu, Arvind Venkataraman, Annsea Park, Benjamin Israelow, Anne L. Wyllie, Chantal B. F. Vogels, M. Catherine Muenker, Arnau Casanovas-Massana, Wade L. Schulz, Joseph Zell, Melissa Campbell, John B. Fournier, Yale IMPACT Research Team, Nathan D. Grubaugh, Shelli Farhadian, Adam V. Wisnewski, Charles Dela Cruz, Saad Omer, Albert I. Ko, Aaron Ring, Akiko Iwasaki

## Abstract

Recent studies have provided insights into innate and adaptive immune dynamics in coronavirus disease 2019 (COVID-19). Yet, the exact feature of antibody responses that governs COVID-19 disease outcomes remain unclear. Here, we analysed humoral immune responses in 209 asymptomatic, mild, moderate and severe COVID-19 patients over time to probe the nature of antibody responses in disease severity and mortality. We observed a correlation between anti-Spike (S) IgG levels, length of hospitalization and clinical parameters associated with worse clinical progression. While high anti-S IgG levels correlated with worse disease severity, such correlation was time-dependent. Deceased patients did not have higher overall humoral response than live discharged patients. However, they mounted a robust, yet delayed response, measured by anti-S, anti-RBD IgG, and neutralizing antibody (NAb) levels, compared to survivors. Delayed seroconversion kinetics correlated with impaired viral control in deceased patients. Finally, while sera from 89% of patients displayed some neutralization capacity during their disease course, NAb generation prior to 14 days of disease onset emerged as a key factor for recovery. These data indicate that COVID-19 mortality does not correlate with the cross-sectional antiviral antibody levels *per se*, but rather with the delayed kinetics of NAb production.

## Main

Coronavirus Disease 2019 (COVID-19) is caused by SARS-CoV-2, which infects host cells via angiotensin-converting enzyme 2 (ACE2) (Hoffmann et al., 2020; Yan et al., 2020). While 80% of infections are mild or asymptomatic (WHO; who.int), moderate and severe COVID-19 patients develop a wide range of symptoms, including respiratory, vascular and neurological complications (Chen et al., 2020a; Huang et al., 2020; Xu et al., 2020). Several studies have linked cellular and humoral immune responses to viral clearance and distinct disease trajectories (Giamarellos-Bourboulis et al., 2020; Huang et al., 2020; Lucas et al., 2020; Rodriguez et al., 2020; Zhou et al., 2020a). For instance, inflammatory cytokines including IFNs, IL-1β, IL-4, IL-6 and IL-18, are associated with worse COVID-19 outcome (Del Valle et al., 2020; Lucas et al., 2020). Importantly, in contrast to the marked decreases in circulating T cells observed in COVID-19 patients (Giamarellos-Bourboulis et al., 2020; Huang et al., 2020; Mathew et al., 2020; Zhou et al., 2020a), circulating B cell do not seem to decrease (Chen et al., 2020a; Lucas et al., 2020). Additionally, several studies reported an overall increase in both IgM and IgG anti-SARS-CoV-2 spike (anti-S IgG), as well as with the presence of neutralizing IgG and IgA antibodies in COVID-19 patients (Barnes et al., 2020; Robbiani et al., 2020; Schmidt et al., 2020; Wang et al., 2020). However, information about how antibody responses affect the course of COVID-19 trajectory, and how they correlate with additional host factors, viral titers and with clinical outcome, is still missing.

### Anti-S and anti-RBD antibodies correlate with distinct COVID-19 outcomes

To profile the SARS-CoV-2-specific humoral immune response, one hundred and seventy-nine hospitalized COVID-19 patients, with a total of 280 samples, were enrolled in this study after admission to Yale-New Haven Hospital (YNHH) between 18 March 2020 and 27 May 2020. In parallel, we enrolled 41 non-hospitalized participants including asymptomatic and mild participants. Additionally, 108 healthcare workers (HCWs) samples served as uninfected healthy controls (SARS-CoV-2-negative by RT–qPCR and serology). Basic demographics and clinical characteristics for each cohort are summarized in Extended Data Table 1. For our initial analysis, hospitalized patients were first stratified based on disease severity into moderate and severe disease groups by supplemental oxygen levels requirements and admission to the intensive care unit (ICU). Further investigations divided COVID-19 patients according to clinical outcomes, stratifying patients that ultimately recovered or die from infection, as previously described (Lucas et al., 2020). The total of 322 samples, collected at hospital admission, include sequential follow-up measurements with a range of one to seven longitudinal time-points per patient that occurred 3–60 days after the onset of symptoms. We assessed viral RNA load, measured by quantitative PCR with reverse transcription (RT–qPCR) using nasopharyngeal swabs; levels of plasma cytokines and chemokines, measured using a Human Cytokine /Chemokine Array 71-403 Plex Panel; leukocyte populations, profiled by flow cytometry using freshly isolated peripheral blood mononuclear cells (PBMCs); and antibodies profile, using both ELISA and neutralizations assays.

Plasma sample analysis showed that 95.1% and 97% of total COVID-19 hospitalized patients had virus-specific IgG against spike (S1) or RBD region of the proteins, respectively, reaching the peak of IgG production around day 15 after symptom onset (Extended Data Fig. 1a-b). The average of anti-S or anti-RBD IgG levels from uninfected control donors, HCW, were used to determine the limit threshold. There were no differences in antibody levels between hospitalized patients of different age, sex or BMI (body mass index) in our cohort (Extended Data Fig. 1c-f). Maximum viral titers from nasopharyngeal swabs were reached approximately 11–13 days after symptom onset. Overall, maximum nasal viral RNA load did not correlate with disease severity; no differences were observed between the non-hospitalized group or moderate and severe patients (Extended Data Fig. 2a). In contrast, hospitalized patients showed a significant increase in maximum anti-S and anti-RBD IgM and IgG levels when compared to non-hospitalized individuals (Figure 1a, b). Additionally, anti-S IgG levels correlated with length of hospitalization among severe, but not moderate patients (Figure 1c). Consistent with this observation, clinical parameters associated with worsened clinical progression, such as intubation, ferritin and D-dimer levels were positively correlated with anti-S IgG levels (Figure 1d, e). In contrast, anti-RBD IgG levels were not correlated with length of hospitalization (Figure 1b-e). Thus, these data indicate that elevated anti-S IgG levels are associated with worse disease outcome in severe COVID-19 patients, confirming previous observations that antibody responses were consistently higher among hospitalized subjects (Chen et al., 2020c; Chen et al., 2020d; Hu et al., 2020; Yu et al., 2020b). Notably, deceased patients did not have higher levels of virus-specific IgG or IgM than the live discharged patients (Fig. 1a). These results indicated a fundamentally different feature of their antibody responses compared to severe disease patients who survived the infection.

**Fig. 1.**
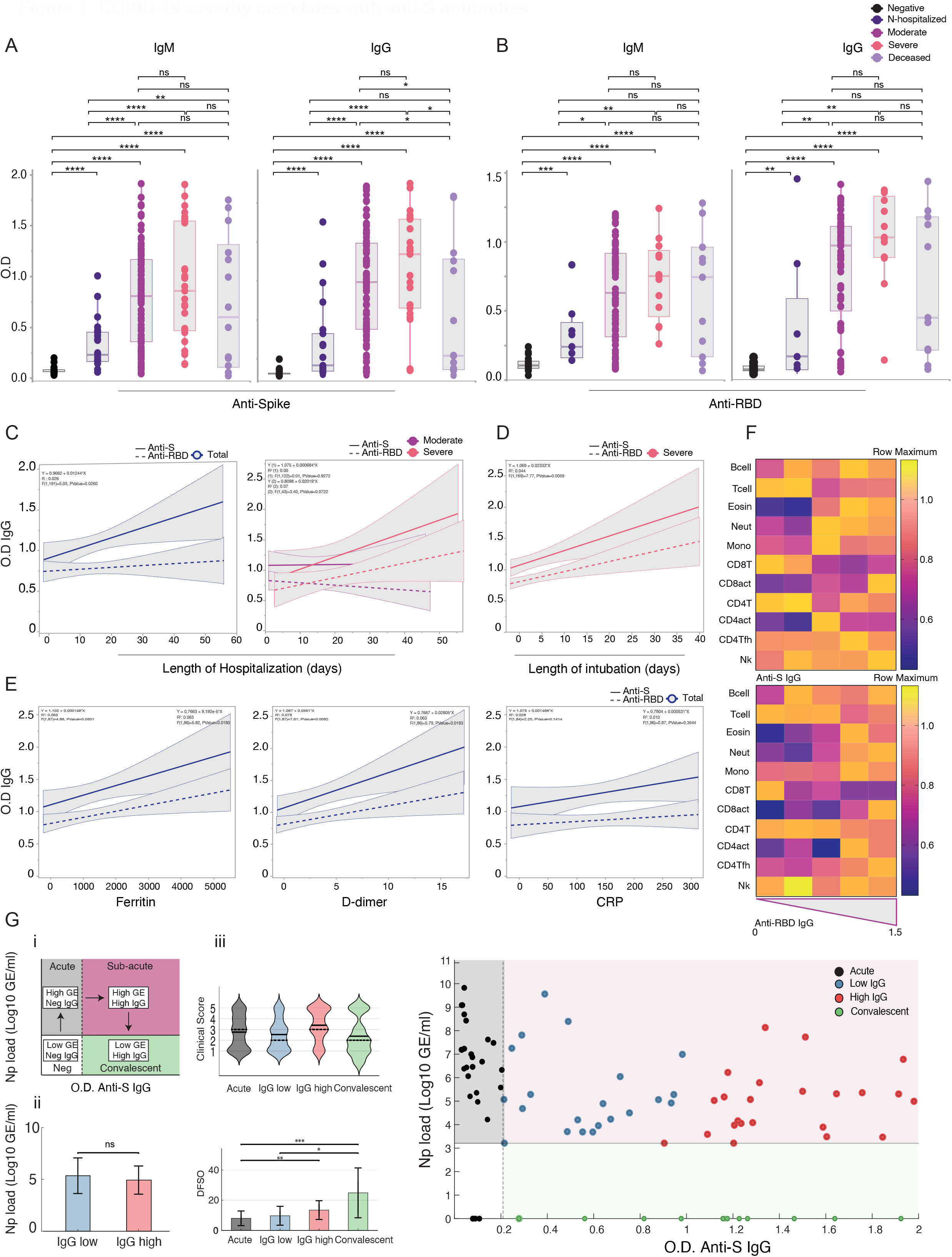
COVID-19 severity correlates with anti-S antibodies. **a-b**, Plasma reactivity to S protein and RBD by ELISA. **a**, Anti-S IgM and IgG comparison in non-hospitalized, moderate, severe and deceased COVID-19 hospitalized patients. IgM (HCW, n = 21; non-hospitalized, n=21; moderate, n = 94; severe, n = 25; deceased, n=14). IgG (HCW, n = 87; non-hospitalized, n=21; moderate, n = 96; severe, n = 23; deceased, n=14). **b**, Anti-RBD IgM and IgG comparison in non-hospitalized, moderate, severe and deceased COVID-19 patients. IgM (HCW, n = 21; non-hospitalized, n=7; moderate, n = 75; severe, n = 13; deceased, n=11). IgG (HCW, n = 21; non-hospitalized, n=6; moderate, n = 74; severe, n = 13; deceased, n=11). Negative controls (HCWs) are shown in black. OD, optical density at 450 nm (OD_450 nm_). N-hospitalized, non-hospitalized. For **a** and **b**, each dot represents a single individual at its maximum antibody titer over the disease course. Horizontal bars indicate mean values. **c**, Correlation and linear regression of virus-specific IgG (OD_450 nm_) and length of hospitalization measured over time. Left, Total patients regardless of disease severity. Regression lines are shown as dark blue, continuous line (Anti -S) or dashed line (Anti-RBD). Right, Patients grouped by disease severity. Regression lines are shown as dark purple (moderate) or pink (severe), continuous line (Anti -S) or dashed line (Anti-RBD). **d-e**, Correlation of virus-specific IgG (OD_450 nm_) and (d) length of intubation and patients’ maximum levels of (e) ferritin, D-dimer, and CRP. **f**, Heat map correlation of the levels of virus-specific IgG (OD_450 nm_) and the major immune cell populations within PBMCs in COVID-19 patients. Subjects are arranged across rows based on their levels of IgG anti-S (upper) or anti-RBD (lower) with each coloured unit indicating the relative distribution of an immune cell population normalized against the same population across all subjects. *K*-means clustering was used to arrange patients and measurements. Eosi, eosinophils. Neut, neutrophils. Mono, monocytes. CD8act, CD4act, CD4Tfh, follicular helper T cells. g, Spectral clustering of COVID-19 patients with S1 IgG+ samples at their first collection time point into IgG low, IgG high, and Convalescent clusters (main, below). Dashed vertical line represents threshold of S1 IgG positivity, and was generated by calculating the 95th percentile of S1 IgG O.D. 450 levels from SARS-CoV-2 negative individuals + 1 standard deviation. Solid horizontal line represents the lower limit of detection for our RT-qPCR assay previously described. (insets, above) i, Graphical schematic of transitions between disease states for COVID-19 patients, colored by disease state (acute, sub-acute, convalescent) ii, Comparison of mean viral loads between Sub-acute clusters using two sample t-test. iii, Violin plots of clinical scores for each cluster. Solid black lines are means of each cluster, whereas dashed lines represent the median value. No significant difference was noted, significance was assessed by Kruskal-Wallis testing corrected for multiple comparisons using Dunn’s method. iv, Days from symptom onset compared for each cluster. Significance was assessed by Kruskal-Wallis testing corrected for multiple comparisons using Dunn’s method. *** p < .001; ** p < .01; *p < .05.

Due to the correlation observed between anti-S IgG levels and disease severity, as well as previous reports describing changes in leukocyte populations in severe COVID-19, including lymphopenia and increased monocytes, neutrophils and eosinophils numbers (Giamarellos-Bourboulis et al., 2020; Huang et al., 2020; Lucas et al., 2020; Mathew et al., 2020; Zhou et al., 2020a) we next assessed whether changes in virus-specific antibodies were linked to alterations in innate and adaptive circulating immune cell types. Anti-S and anti-RBD IgG levels negatively correlated with T cells and positively with monocytes, neutrophils and eosinophils numbers, but no correlation was found with circulating NK or B cells (Figure 1f). Furthermore, we observed a positive correlation between anti-RBD, but not anti-S, IgG levels and circulating T follicular helper CD4^+^ T cells (Tfh) as well as CD38+HLA-DR+ TCR-activated CD4 T cells (CD4act) (Figure 1f). Further stratification of hospitalized patients at their initial collection time point into acute, sub-acute and convalescent phases of COVID-19, indicated a correlation between anti-S IgG and clinical disease severity, even among patients with equivalent viral loads in the same phase of disease onset (Figure 1g). Specifically, between clusters matched for equivalent viral loads and similar days from symptom onset, we noted an increase in the mean clinical score of the high S IgG group relative to the low S IgG group, although this was not statistically significant. Importantly, it was clear that the development of anti-S IgG responses did not correlate with general improvements in patients clinical scores. Given these results, we conclude that anti-S IgG antibodies positively correlate with COVID-19 severity and appear to offer a limited ability to modify disease trajectory once developed during natural SARS-CoV-2 infection. We conclude that anti-S IgG antibodies positively correlate with COVID-19 severity, along with the circulating levels of monocytes, neutrophils and eosinophils, but independent of circulating T cells, Tfh or viral load.

### Delayed antibody production in lethal COVID-19

Given the lower levels of antiviral antibodies found in deceased patients, we next addressed whether the timing of antibody responses differ between severe vs. lethal disease. Longitudinal analysis revealed distinct kinetics: discharged patients reached an early peak of anti-S and anti-RBD IgG levels faster than deceased patients (Figure 2a). In contrast, deceased patients reached higher maximum levels of anti-S IgM and IgG than discharged in later stages of disease (Figure 2a). Longitudinal antibody trajectories between discharged and deceased groups were consistent with their distinct capacity to clear the virus; i.e. discharged patients were more efficient in viral clearance when compared side-by-side with deceased patients (Figure 2b). Additionally, lower levels of nasal viral RNA load measured at the time of maximum antibody levels were observed in discharged patients (Figure 2c). We did not observe differences in B cell dynamics in patients with distinct clinical outcomes (Extended Data Fig. 3a, b). Despite no differences between discharged and deceased groups at aggregate levels, longitudinal analysis indicated a higher frequency of Tfh at DfSO 10-15 in discharged than in deceased COVID-19. Thus, death from COVID-19 correlated with a delay in the development of virus-specific IgG and virus clearance.

**Fig. 2.**
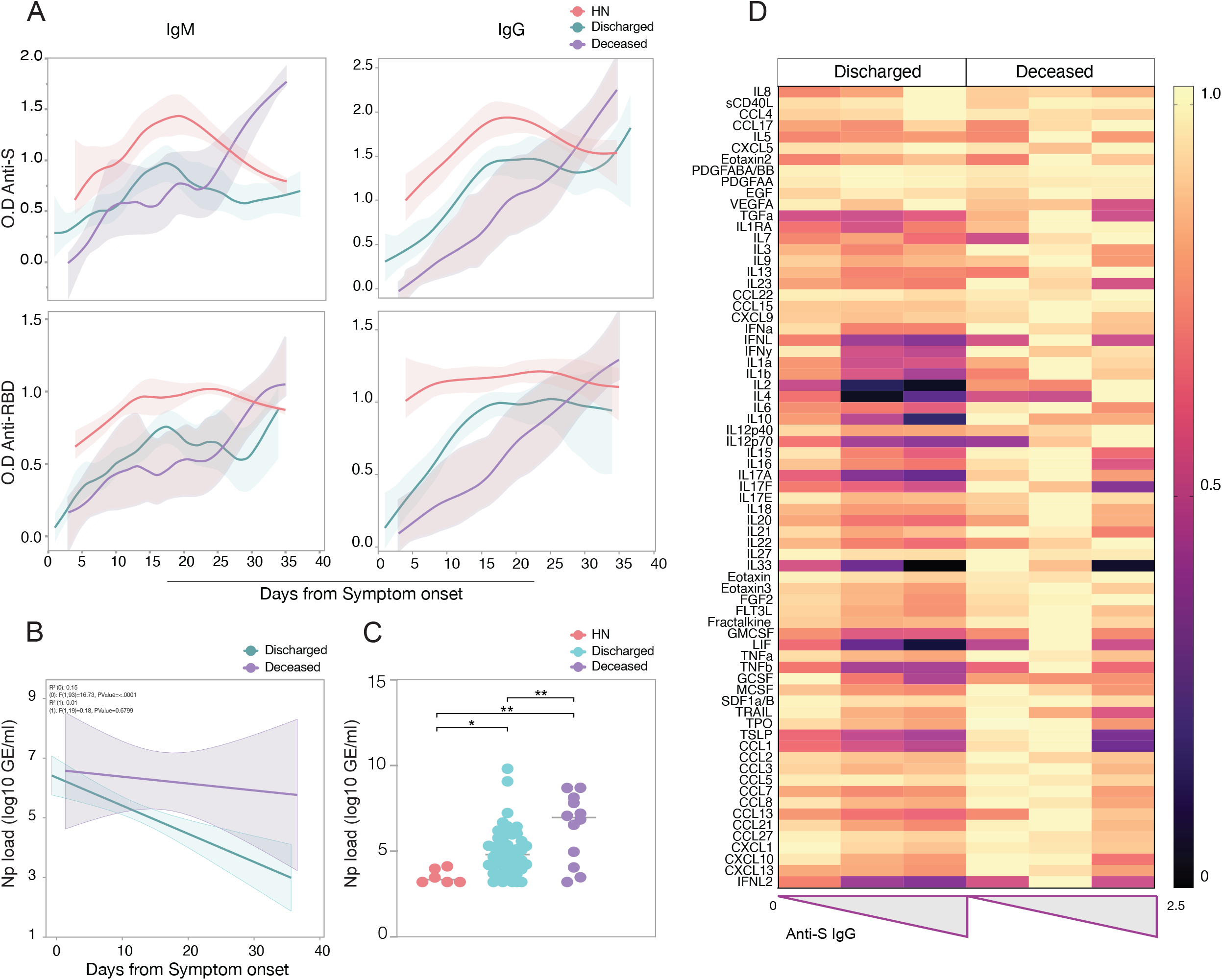
Serum antibody kinetics reveals distinct COVID-19 outcomes. **a**, Patients plasma reactivity to S protein and RBD by ELISA. Anti-S and Anti-RBD IgM and IgG comparison in discharged or deceased patients. Longitudinal data plotted over time continuously. Regression lines are shown as light blue (discharged), purple (deceased) and red (High neutralizers). Shading represents 95% CI and are coloured accordingly. Anti-S IgM, (Discharged, n=127; Deceased, n=14). Anti-S IgG, (Discharged, n=128; Deceased, n=14). Anti-RBD IgM, (Discharged, n=88; Deceased, n=11). Anti-S RBD, (Discharged, n=87; Deceased, n=11). **b-c**, Viral loads measured by nasopharyngeal swabs are plotted as log_10_ of genome equivalents (GE). b, Viral loads against time after symptom onset accordingly with patient’s outcome. **b**, Regression lines are shown as light blue (discharged) or purple (deceased). **c**, Viral load measured in discharged, deceased and high neutralizer patients. (HN, n=6; discharged, n = 53; deceased, n = 12). Each dot represents the viral load of a single individual at its maximum antibody titer over the disease course. **d**, Heat map correlation of the levels of Anti-S IgG (OD_450 nm_) and plasma cytokines/chemokines measurements within discharged (n=146) or deceased patients (n=26). Subjects are arranged across rows based on their levels of anti-S IgG with each coloured unit indicating the relative cytokine concentration (log_10)_ normalized against the same population across all subjects. *K*-means clustering was used to arrange patients and measurements.

We next assessed a possible correlation between cytokines and chemokines levels and virus-specific antibody production. Discharged patients showed a positive correlations between anti-S IgG and several chemokines, growth factors and tissue repair mediators, including sCD40L, CXCL5, IL-8, VEGFA, CCL4, CCL17, Eotaxin2 (Figure 2d); consistent with a “protective signature” we recently observed in these patients, who recover from COVID-19 (Lucas et al., 2020). Additionally, discharged patients showed a negative correlation between anti-S IgG and plasma inflammatory markers previously associated with poor disease outcomes and death, such as IFN-I, II and III, IL-1, IL-6, IL-17 and IL-10 (Figure 2d). Deceased patients in contrast showed fewer correlations with anti-S IgG levels and overall heightened levels of plasma cytokines and chemokines (Figure 2d).

### Early production of neutralizing antibodies correlates with COVID-19 recovery

Production of anti-S/RBD IgG antibodies is generally associated with virus neutralization (Robbiani et al., 2020), and have been linked with protection against SARS-CoV-2 infection after vaccination in animal models (Folegatti et al., 2020; Yu et al., 2020a). We next assessed the kinetics of neutralizing antibodies (NAb) produced against SARS-CoV-2 by performing a neutralization assay using wild-type SARS-CoV-2 in patients that reached high levels of anti-S IgG (O.D.>1.2) at any point during their disease course. In addition, all deceased patients were included in the analysis regardless of whether they reached the limit cutoff point; samples from health care workers negative for SARS-CoV-2 RT-qPCR were used as controls samples and were below the threshold for anti-S/RBD ELISA. Previous studies have reported a low frequency of convalescent COVID-19 patients showing potent neutralization capacity of over 1:1000 titers (Robbiani et al., 2020). Indeed, while 89% of patients in our cohort showed some neutralization capacity during their disease course, 80-86% of hospitalized patients exhibited neutralizing activity only at lower dilutions (1:10-1:90 titers), and only 7-27% of patients showed neutralizing activity at higher dilutions (1:810-1:2430 titers) (Figure 3a, b and Extended Data Fig. 4a, b). The neutralization levels were lower among non-hospitalized participants with mild disease, even at lower dilutions of 1:90 (31%) and 1:270 (4%) titers (Extended Data Fig. 4b). Based on this stratification, we designated patients with >1:810 neutralizing titer as high neutralizers (HN). The half-maximal neutralizing titer (PRNT50) was undetectable for 15.6% of patients, while 25.9% of patients had PRNT50 at 1:270 and only 1.3% of patient at 1:810. Of note, the levels of PRNT50 were not significantly different between hospitalized patients stratified by disease severity and outcome (Figure 3c, d). These PRNT50 patterns among hospitalized patients are consistent with a recent pre-print (Garcia-Beltran et al., 2020).

**Fig. 3.**
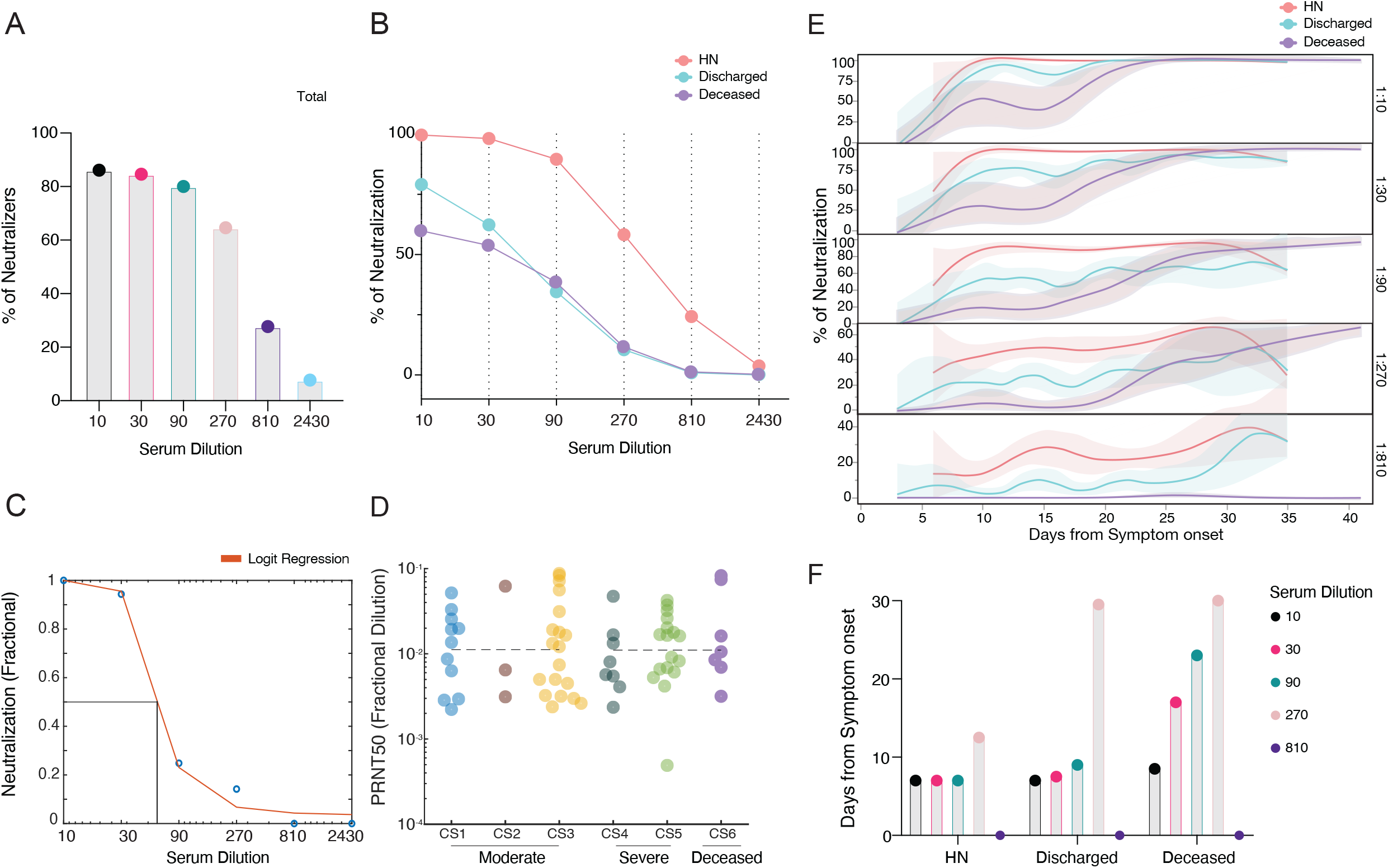
Neutralizing antibodies temporal dynamics distinguishes discharged and deceased COVID-19 patients. **a-e**, Longitudinal neutralization assay using wild-type SARS-CoV-2. **a**, Frequency of neutralizers among patients with high anti-S IgG levels (O.D.>1,2), n=65. **b**, Neutralization capacity between discharged (light blue), deceased (purple) and high neutralizers (red) at the experimental sixfold serially dilutions (from 1:3 to 1:2430). Health care workers, below the threshold for anti-S/RBD ELISA, were used as negative controls. HN, high neutralizers. (HCW, n = 22; discharged, n=35; deceased, n=17; high neutralizers, n=13). **c-d**, Logistic Regression and neutralization titer (PRNT50) according to clinical severity scale as described in Methods. CS, clinical score. **e**, Longitudinal data plotted over time of neutralization capacity between discharged (light blue), deceased (purple) and high neutralizers (red) at the experimental sixfold serially dilutions (from 1:3 to 1:2430). **f**, Average of days from symptom onset to reach 50% of neutralization at each experimental serum dilution between groups.

Importantly, our longitudinal analysis also revealed faster NAb kinetics, as well as higher peak, in discharged than decease patients. Discharged patients reached 50% of neutralization at 1:90 titer around day 9 post symptoms onset, while deceased patients peaked 1 and 2 weeks later in 1:30 and 1:90 titers, respectively (Figure 3e, f). We then compared maximum anti-S, anti-RBD IgG titers and viral load between high neutralizers, discharged and deceased patients, when each group reached 50% of neutralization. As expected, the levels of anti-S/RBD IgG antibodies strongly correlated with PRNT50, and no significant differences were observed between discharged and deceased groups (Extended Data Fig. 4c). Despite equivalent maximum or PRNT50 NAb titers, distinct temporal antibody dynamics was strongly linked with clinical disease outcome. Remarkably, the high neutralizers had maximum levels of anti-RBD IgG and NAb from the very first sampling (5 days post disease onset), and maintained high levels throughout the hospital stay (Figure 2b). High neutralizers presented lower levels of nasal viral RNA load measured at the time of maximum antibody level in comparison to deceased or discharged patients (Figure 2c).

Finally, we asked whether the timing of NAb production dictates disease trajectory. Patients were grouped into those who developed >50% neutralization activity at 1:90 titer NAb levels before 14 days of symptoms onset (early), and those who did not (late) (Figure 4a). Among patients with high levels of Anti-S IgG (OD>1.2), 48.2% had >50% of neutralization activity at 1:90 titer, but only 17.8% at 1:270 (Figure 4b). Early neutralization activity did not correlate with age or BMI and the frequency of males and females were not significantly different between early or late neutralizers (Figure 4c). Nevertheless, early NAb production correlated with improving clinical signs and lower mortality than late neutralizers, which overall showed worst disease progression and higher mortality (Figure 4d, e). Moreover, the maximum viral loads reached over the disease course were lower in early neutralizers (Figure 4f). Together, these data indicate that clinical trajectories and outcomes do not correlate with cross-sectional levels of NAb production *per se*, but rather with the timing and kinetics of NAb production.

**Fig. 4.**
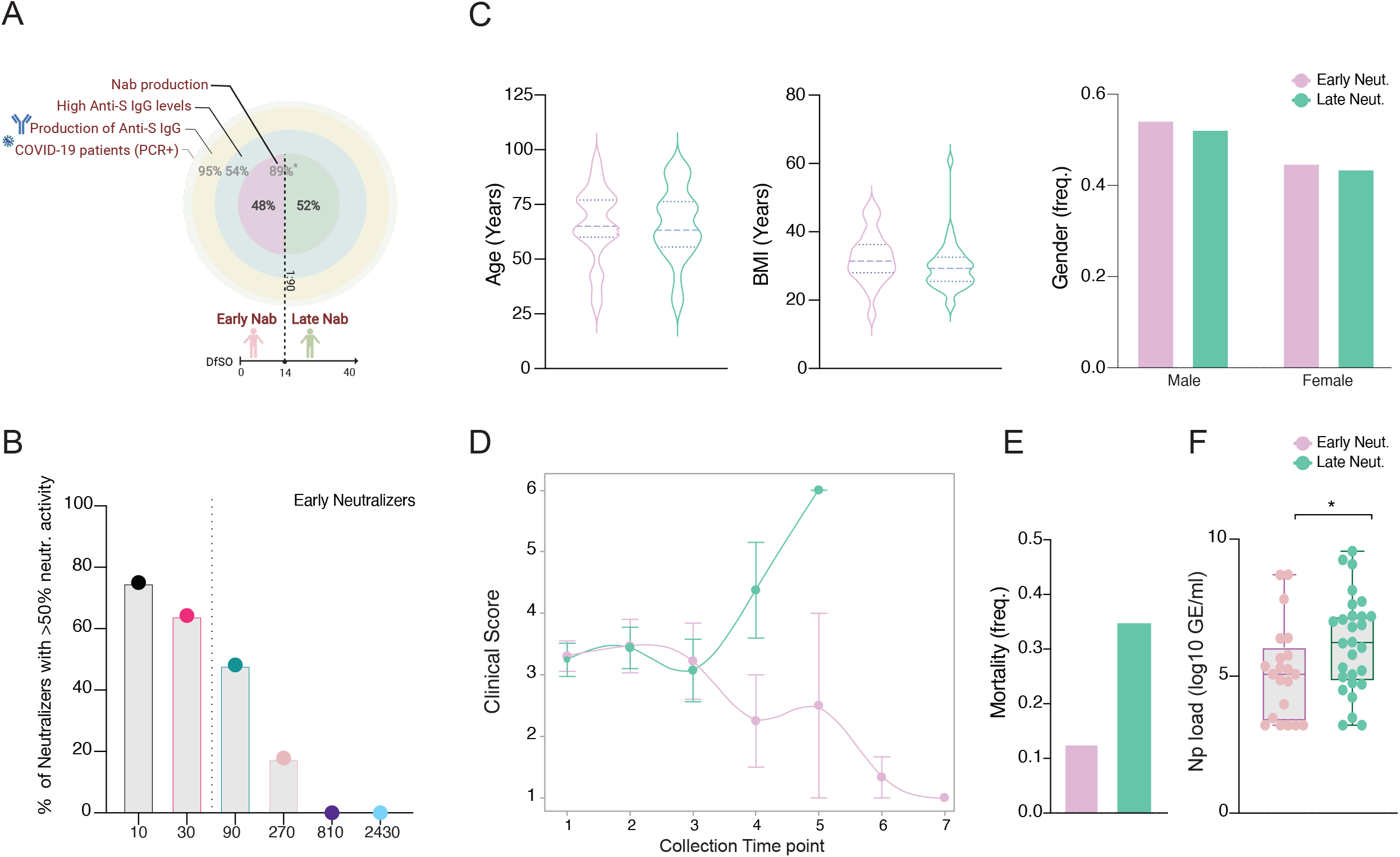
Early neutralizing antibodies correlates with better COVID-19 clinical trajectory. Patients stratification by early NAb capacity, based on levels of Anti-S IgG, PRNT50 titers and days from symptom onset. **a**, Cohort overview by IgG Anti-S titers (3 external circles) and NAb production (internal circle). Frequency of patients in each level are indicated in light grey. Frequency of early and late neutralizers stratified based on days from symptom onset at 1:90 dilution is indicated in black. NAb, neutralizing antibodies. DfSO, days from symptom onset. * Frequency of patients with NAb capacity over the disease. course within patients with high levels of anti-S IgG. **b**, Frequency of neutralizers with >50% neutralization activity until 14 days after symptom onset. **c**, Distribution of age, BMI and frequency of males and females among early (>50% neutralization activity in 1:90 titer before day 14 after symptom onset) or late (<50% neutralization activity in 1:90 titer before day 14 after symptom onset) neutralizers length, as determined in (a). **d**, Disease progression measured by clinical severity score for patients in each group. Data (mean ± s.e.m) are ordered by the collection time points for each patient, with regular collection intervals of 3–4 days. **e**, Percentage of mortality in each group. (Early neutralizers, n=24; Late neutralizers, n=38). **f**, Viral load measured by nasopharyngeal swabs plotted as log10 of genome equivalents in early and late neutralizers. (Early neutralizers, n=21; Late neutralizers, n=28). Each dot represents a single individual at its maximum antibody titer over the disease course. Box analysis with minimum and maximum represented for each group.

## Discussion

The nature of virus-specific serum antibodies evolves rapidly following infection and has been previously correlated with disease severity in patients infected with different coronaviruses, including MERS, SARS-CoV-1 and SARS-CoV-2 (Chen et al., 2020c; Chen et al., 2020d; Hu et al., 2020; Yu et al., 2020b). Our analyses not only confirm a link between SARS-CoV-2-specific antibody responses and COVID-19 severity, but also suggest a specific time-window in which neutralizing antibodies must develop in order to improve viral control and diseases outcomes.

Our temporal analyses revealed a distinct antibody kinetics between discharged and deceased patients. Deceased patients showed slower antibody dynamics even though they reached higher levels later in the disease trajectory. A recent study performing longitudinal analysis also reported a delayed and incomplete humoral immune response in COVID-19 deceased patients (Zohar et al., 2020). Our study confirms and extends these observations by demonstrating that seroconversion kinetics between discharged and deceased patients can directly impact viral clearance, with faster viral control or prolonged viral shedding, respectively. Additionally, our observations that discharged patients show a negative correlation between anti-S IgG and plasma inflammatory markers uncovers a potential role of antibodies in protecting these patients from immunopathology, especially in light of our early study describing a severe inflammatory signature in COVID-19 patients (Lucas et al., 2020). Somehow, this protective role of antiviral antibodies fail to operate in deceased patients, as we saw loss of correlation between anti-S IgG and protective tissue repair growth factors. Our data suggest that the loss of protective role of antibodies in lethal disease is due to their late onset.

Our study demonstrated that neutralizing antibody responses developed within 14 days of symptom onset correlated with recovery, while those induced at later timepoints appear to lose this protective effect. It is unclear why antibodies generated after this time point are unable to promote viral clearance and recovery in COVID-19 patients. We speculate that the virus might become inaccessible to the antibodies after a certain time point, by establishing infection within immune privileged tissues. Alternatively, disease may be driven by late-onset antibody-mediated immunopathology. For instance, antibodies from severe COVID-19 patients show pro-inflammatory Fc modifications signature, including high levels of afucosylated IgG1 (Abry et al., 2020), which could potentially drive pathologic responses. Consistent with this notion is our finding that anti-S but not anti-RBD antibody levels in COVID-19 patients correlate with disease severity, length of hospital stay, length of intubation and various clinical parameters of disease. In addition, the levels of anti-S IgG, when matched for similar viral load and days from symptom onset, correlated with COVID-19 severity. Future studies are needed to address the precise mechanism of the failure of late antibody responses, and the potential immunopathological roles of anti-S IgG.

Recent *post mortem* tissue analysis from patients with lethal SARS-CoV-2 infection suggested a defective induction of germinal centers, including inefficient generation of Tfh cells (Kaneko et al., 2020); data consistent with our observations. Nevertheless, because our analysis did not include isolation of secondary lymphoid tissues, we were unable to assess the dynamics of B cell populations in lymph nodes and may explain why we failed to observe differences in B cell dynamics despite our observations of general differences in the kinetics of humoral responses across patient cohorts. We observed a positive correlation between anti-S IgG and Tfh in discharged but not deceased patients.

The use of convalescent plasma has been proposed as an urgent therapeutic modality for COVID-19 patients. However, large clinical trials conducted to evaluate the efficacy of plasma therapy failed to observe clear benefits (Agarwal et al., 2020; Holliman et al., 1987; Pathak, 2020; Valk et al., 2020; Zhou et al., 2020b), perhaps due to variable amount of neutralizing capacity in the donor pool. Potent neutralizing monoclonal antibodies cloned from B cells isolated from convalescent individuals were proposed to address this issue in COVID-19 therapeutics. However, a recent large trial of monoclonal antibodies also failed to observe improvements when administered to late stage severe or hospitalized patients (Eli Lily); similar observations were made with trials by Regeneron (Lilly statement; Regeneron statement; FDA; fda.gov). Recently, additional early trials have shown promising results in people who were newly infected with the virus (Chen et al., 2020b). Our results demonstrated that anti-S or neutralizing antibodies observed within the 14 days of symptom onset correlated with improved disease trajectory. These data suggest that antibody-based therapies may benefit patients most when given within this two-week time window.

## METHODS

### Ethics statement

This study was approved by Yale Human Research Protection Program Institutional Review Boards (FWA00002571, protocol ID 2000027690). Informed consent was obtained from all enrolled patients and healthcare workers.

### Patients

One hundred and seventy-nine patients admitted to YNHH with COVID-19 between 18 March 2020 and 27 May 2020 were included in this study. Additionally, forty-one non hospitalized participants were enrolled in this study, including asymptomatic and mild individuals. This participants were enrolled by the IMPACT group (25) and an additional 16 serum samples from mild COVID-19 cases from the Connecticut National Guard. HCW participants, screened serially (every two weeks) served as uninfected healthy controls (SARS-CoV-2-negative by RT–qPCR and serology). No statistical methods were used to predetermine sample size. Patients were scored for COVID-19 disease severity through review of the electronic health records (EMR) at each longitudinal time point. Scores were assigned by a clinical infectious disease physician according to a custom-developed disease severity scale. Moderate disease status (clinical score 1–3) was defined as: SARS-CoV-2 infection requiring hospitalization without supplementary oxygen (1); infection requiring non-invasive supplementary oxygen (<3 l/min to maintain SpO_2_ >92%) (2); and infection requiring non-invasive supplementary oxygen (>3 l/min to maintain SpO_2_ >92%, or >2 l/min to maintain SpO_2_ >92% and had a high-sensitivity C-reactive protein (CRP) >70) and received tocilizumab). Severe disease status (clinical score 4 or 5) was defined as infection meeting all criteria for clinical score 3 and also requiring admission to the ICU and >6 l/min supplementary oxygen to maintain SpO_2_ >92% (4); or infection requiring invasive mechanical ventilation or extracorporeal membrane oxygenation (ECMO) in addition to glucocorticoid or vasopressor administration (5). Clinical score 6 was assigned for deceased patients. For all patients, days from symptom onset were estimated as follows: (1) highest priority was given to explicit onset dates provided by patients; (2) next highest priority was given to the earliest reported symptom by a patient; and (3) in the absence of direct information regarding symptom onset, we estimated a date through manual assessment of the EHR by an independent clinician. Demographic information was aggregated through a systematic and retrospective review of the EHR and was used to construct Extended Data Table 1. Symptom onset and aetiology were recorded through standardized interviews with patients or patient surrogates upon enrolment in our study, or alternatively through manual EHR review if no interview was possible owing to clinical status. The clinical data were collected using EPIC EHR and REDCap 9.3.6 software. At the time of sample acquisition and processing, investigators were completely unaware of the patients’ conditions. Blood acquisition was performed and recorded by a separate team. Information of patients’ conditions was not available until after processing and analysing raw data by flow cytometry and ELISA. A clinical team, separate from the experimental team, performed chart reviews to determine patients’ relevant statistics. Cytokines and flow cytometry analyses were blinded. Patients clinical information and clinical scores coding were only revealed after data collection.

### Isolation of patient plasma and PBMCs

Whole blood was collected in sodium heparin-coated vacutainers and kept on gentle agitation until processing. All blood was processed on the day of collection. Plasma samples were collected after centrifugation of whole blood at 400*g* for 10 min at room temperature (RT) without brake. The undiluted serum was then transferred to 15-ml polypropylene conical tubes, and aliquoted and stored at −80 °C for subsequent analysis. PBMCs were isolated using Histopaque (Sigma-Aldrich, #10771-500ML) density gradient centrifugation in a biosafety level 2+ facility. After isolation of undiluted serum, blood was diluted 1:1 in room temperature PBS, layered over Histopaque in a SepMate tube (StemCell Technologies; #85460) and centrifuged for 10 min at 1,200*g*. The PBMC layer was isolated according to the manufacturer’s instructions. Cells were washed twice with PBS before counting. Pelleted cells were briefly treated with ACK lysis buffer for 2 min and then counted. Percentage viability was estimated using standard Trypan blue staining and an automated cell counter (Thermo-Fisher, #AMQAX1000).

### SARS-CoV-2 specific-antibody measurements

ELISAs were performed as previously described (Amanat et al., 2020). In short, Triton X-100 and RNase A were added to serum samples at final concentrations of 0.5% and 0.5mg/ml respectively and incubated at room temperature (RT) for 30 minutes before use to reduce risk from any potential virus in serum. 96-well MaxiSorp plates (Thermo Scientific #442404) were coated with 50 μl/well of recombinant SARS Cov-2 S1 protein (ACROBiosystems #S1N-C52H3-100ug) at a concentration of 2 μg/ml in PBS and were incubated overnight at 4 °C. The coating buffer was removed, and plates were incubated for 1h at RT with 200 μl of blocking solution (PBS with 0.1% Tween-20, 3% milk powder). Serum was diluted 1:50 in dilution solution (PBS with 0.1% Tween-20, 1% milk powder) and 100 μl of diluted serum was added for two hours at RT. Plates were washed three times with PBS-T (PBS with 0.1% Tween-20) and 50 μl of HRP anti-Human IgG Antibody (GenScript #A00166, 1:5,000) or anti-Human IgM-Peroxidase Antibody (Sigma-Aldrich #A6907, 1:5,000) diluted in dilution solution added to each well. After 1 h of incubation at RT, plates were washed three times with PBS-T. Plates were developed with 100 μl of TMB Substrate Reagent Set (BD Biosciences #555214) and the reaction was stopped after 15 min by the addition of 2 N sulfuric acid. Plates were then read at a wavelength of 450 nm and 570nm.

### Cytokine and chemokine measurements

Patient serum was isolated as before and aliquots were stored at −80 °C. Sera were shipped to Eve Technologies (Calgary, Alberta, Canada) on dry ice, and levels of cytokines and chemokines were measured using the Human Cytokine Array/Chemokine Array 71-403 Plex Panel (HD71). All samples were measured upon the first thaw.

### Viral RNA measurements

Nasopharyngeal swabs samples were collected approximately every four days, for SARS-CoV-2 RT–qPCR analysis where clinically feasible. RNA concentrations were measured as previously described (Wyllie et al., 2020). In brief, total nucleic acid was extracted from 300 μl of viral transport medium (nasopharyngeal swab) using the MagMAX Viral/Pathogen Nucleic Acid Isolation kit (ThermoFisher Scientific) with a modified protocol and eluted into 75 μl elution buffer. We used 5 μl of extracted nucleic acid as template in a RT-qPCR assay to detect SARS-CoV-2 RNA (Vogels et al., 2020), using the US CDC real-time RT–qPCR primer/probe sets for 2019-nCoV_N1, 2019-nCoV_N2, and the human RNase P (RP) as an extraction control. Virus RNA copies were quantified using a tenfold dilution standard curve of RNA transcripts that we previously generated (Vogels et al., 2020). The lower limit of detection for SARS-CoV-2 genomes assayed by qPCR in nasopharyngeal specimens was established as described (Vogels et al., 2020). In addition to a technical detection threshold, we also used a clinical referral threshold (detection limit) to either: (1) refer asymptomatic HCWs for diagnostic testing at a CLIA-approved laboratory; or (2) cross-validate results from a CLIA-approved laboratory for SARS-CoV-2 qPCR-positive individuals upon study enrolment. Individuals above the technical detection threshold, but below the clinical referral threshold, were considered SARS-CoV-2 positive for the purposes of our research.

### Flow cytometry

Antibody clones and vendors were as follows: BB515 anti-hHLA-DR (G46-6) (1:400) (BD Biosciences), BV785 anti-hCD16 (3G8) (1:100) (BioLegend), PE-Cy7 anti-hCD14 (HCD14) (1:300) (BioLegend), BV605 anti-hCD3 (UCHT1) (1:300) (BioLegend), BV711 anti-hCD19 (SJ25C1) (1:300) (BD Biosciences), AlexaFluor647 anti-hCD1c (L161) (1:150) (BioLegend), biotin anti-hCD141 (M80) (1:150) (BioLegend), PE-Dazzle594 anti-hCD56 (HCD56) (1:300) (BioLegend), PE anti-hCD304 (12C2) (1:300) (BioLegend), APCFire750 anti-hCD11b (ICRF44) (1:100) (BioLegend), PerCP/Cy5.5 anti-hCD66b (G10F5) (1:200) (BD Biosciences), BV785 anti-hCD4 (SK3) (1:200) (BioLegend), APCFire750 or PE-Cy7 or BV711 anti-hCD8 (SK1) (1:200) (BioLegend), BV421 anti-hCCR7 (G043H7) (1:50) (BioLegend), AlexaFluor 700 anti-hCD45RA (HI100) (1:200) (BD Biosciences), PE anti-hPD1 (EH12.2H7) (1:200) (BioLegend), APC anti-hTIM3 (F38-2E2) (1:50) (BioLegend), BV711 anti-hCD38 (HIT2) (1:200) (BioLegend), BB700 anti-hCXCR5 (RF8B2) (1:50) (BD Biosciences), PE-Cy7 anti-hCD127 (HIL-7R-M21) (1:50) (BioLegend), PE-CF594 anti-hCD25 (BC96) (1:200) (BD Biosciences), BV711 anti-hCD127 (HIL-7R-M21) (1:50) (BD Biosciences), BV421 anti-hIL17a (N49-653) (1:100) (BD Biosciences), AlexaFluor 700 anti-hTNFa (MAb11) (1:100) (BioLegend), PE or APC/Fire750 anti-hIFNy (4S.B3) (1:60) (BioLegend), FITC anti-hGranzymeB (GB11) (1:200) (BioLegend), AlexaFluor 647 anti-hIL-4 (8D4-8) (1:100) (BioLegend), BB700 anti-hCD183/CXCR3 (1C6/CXCR3) (1:100) (BD Biosciences), PE-Cy7 anti-hIL-6 (MQ2-13A5) (1:50) (BioLegend), PE anti-hIL-2 (5344.111) (1:50) (BD Biosciences), BV785 anti-hCD19 (SJ25C1) (1:300) (BioLegend), BV421 anti-hCD138 (MI15) (1:300) (BioLegend), AlexaFluor700 anti-hCD20 (2H7) (1:200) (BioLegend), AlexaFluor 647 anti-hCD27 (M-T271) (1:350) (BioLegend), PE/Dazzle594 anti-hIgD (IA6-2) (1:400) (BioLegend), PE-Cy7 anti-hCD86 (IT2.2) (1:100) (BioLegend), APC/Fire750 anti-hIgM (MHM-88) (1:250) (BioLegend), BV605 anti-hCD24 (ML5) (1:200) (BioLegend), BV421 anti-hCD10 (HI10a) (1:200) (BioLegend), BV421 anti-CDh15 (SSEA-1) (1:200) (BioLegend), AlexaFluor 700 Streptavidin (1:300) (ThermoFisher), BV605 Streptavidin (1:300) (BioLegend). In brief, freshly isolated PBMCs were plated at 1–2 × 10^6^ cells per well in a 96-well U-bottom plate. Cells were resuspended in Live/Dead Fixable Aqua (ThermoFisher) for 20 min at 4 °C. Following a wash, cells were blocked with Human TruStan FcX (BioLegend) for 10 min at RT. Cocktails of desired staining antibodies were added directly to this mixture for 30 min at RT. For secondary stains, cells were first washed and supernatant aspirated; then to each cell pellet a cocktail of secondary markers was added for 30 min at 4 °C. Prior to analysis, cells were washed and resuspended in 100 μl 4% PFA for 30 min at 4 °C. Following this incubation, cells were washed and prepared for analysis on an Attune NXT (ThermoFisher). Data were analysed using FlowJo software version 10.6 software (Tree Star). The specific sets of markers used to identify each subset of cells are summarized in Extended Data Fig.5.

### Cell lines and virus

VeroE6 kidney epithelial cells) were cultured in Dulbecco’s Modified Eagle Medium (DMEM) supplemented with 1% sodium pyruvate (NEAA) and 5% fetal bovine serum (FBS) at 37°C and 5% CO2. The cell line was obtained from the ATCC and has been tested negative for contamination with mycoplasma. SARS-CoV-2, strain USA-WA1/2020, was obtained from BEI Resources (#NR-52281) and was amplified in VeroE6 cells. Cells were infected at a MOI 0.01 for four three days to generate a working stock and after incubation the supernatant was clarified by centrifugation (450g × 5min) and filtered through a 0.45-micron filter. The pelleted virus was then resuspended in PBS then aliquoted for storage at − 80°C. Viral titers were measured by standard plaque assay using Vero E6 cells. Briefly, 300 µl of serial fold virus dilutions were used to infect Vero E6 cells in MEM supplemented NaHCO3, 4% FBS 0.6% Avicel RC-581. Plaques were resolved at 48h post infection by fixing in 10% formaldehyde for 1h followed by with 0.5% crystal violet in 20% ethanol staining. Plates were rinsed in water to plaques enumeration. All experiments were performed in a biosafety level 3 with the Yale Environmental Health and Safety office approval.

### Neutralization assay

Patients and healthy donors sera were isolated as before and then heat treated for 30m at 56 °C. Sixfold serially diluted plasma, from 1:3 to 1:2430 were incubated with SARS-CoV-2 for 1 h at 37 °C. The mixture was subsequently incubated with Vero E6 cells in a 6-well plate for 1h, for adsorption. Then, cells were overlayed with MEM supplemented NaHCO_3_, 4% FBS 0.6% Avicel mixture. Plaques were resolved at 40h post infection by fixing in 10% formaldehyde for 1h followed by staining in 0.5% crystal violet. All experiments were performed in parallel with negative controls sera, in a establish viral concentration to generate 60-120 plaques/well.

### Statistical analysis

All analysis of patient samples was conducted using Matlab 2020a, GraphPad Prism 9, and JMP 15. Patient heatmaps were clustered using the K-means algorithm. For Figure 1g, patient samples with positive IgG values were z-scored and subsequently clustered using a spectral clustering technique with a ‘cityblock’ distance metric (Matlab 2020a). Multiple group comparisons were analyzed by running both parametric (ANOVA) and non-parametric (Kruskal–Wallis) statistical tests, as indicated in figure legends. For the comparison between stable groups, two-sided unpaired t-test was used for the comparison.

## Data Availability

All the background information on HCWs, clinical information for patients, and raw data used in this study are included in a Supplementary Table 1. Additionally all of the raw fcs files for the flow cytometry analysis are available.

## Data availability

All the background information on HCWs, clinical information for patients, and raw data used in this study are included in a Supplementary Table 1. Additionally, all of the raw fcs files for the flow cytometry analysis are available at (not yet available).

## Acknowledgements

We thank M. Linehan for technical and logistical assistance, and C. Wilen and D. Mucida for discussions. We also thank C. Wilen for kindly providing the virus. This work was supported by the Women’s Health Research at Yale Pilot Project Program (A.I.) [Fast Grant from Emergent Ventures at the Mercatus Center, Mathers Foundation, and the Ludwig Family Foundation, the Department of Internal Medicine at the Yale School of Medicine, Yale School of Public Health and the Beatrice Kleinberg Neuwirth Fund. IMPACT received support from the Yale COVID-19 Research Resource Fund. A.I. is an Investigator of the Howard Hughes Medical Institute. C.L. is a Pew Latin American Fellow. P.Y is supported by Gruber Foundation and the NSF. B.I. is supported by NIAID 2T32AI007517-16. C.B.F.V. is supported by NWO Rubicon 019.181EN.004.

## Author contributions

A.I.K. and A.I. conceived the study. C.L., J.K., J.S., J.E.O. and T.M collected and processed patient PBMC and plasma samples. C.L. and J.K., performed the neutralization assays and data analyses. P.W performed the flow cytometry and C.L. analysed the flow data. J.S., and B.I. collected epidemiological and clinical data. F.L. performed the SARR-CoV-2 specific antibody ELISAs. A.R. supervised the ELISAs. A.L.W., C.B.F., P.L., A.V., A.P., and M.T. performed samples processing, extractions, and RT-qPCR assays, under supervision of N.D.G. A.C.-M., M.C.M processed and stored patient specimens. J.Z. and A.V.W assisted in mild disease volunteer recruitment. M.C., J.B.F., C.D.C. and S.F. assisted hospitalized patients’ identification and enrolment. W.L.S. supervised clinical data management. C.L. and A.I. drafted the manuscript. All authors helped to edit the manuscript. A.I. secured funds and supervised the project.

## Competing interests

AI served as a consultant for Spring Discovery and Adaptive Biotechnologies.

## Yale IMPACT Research Team

Abeer Obaid^10^, Adam J. Moore^2^, Alice Lu-Culligan^1^, Allison Nelson^10^, Anderson Brito^2^, Angela Nunez^10^, Anjelica Martin^1^, Anne E. Watkins^2^, Bertie Geng^10^, Chaney C. Kalinich^2^, Christina A. Harden^2^, Codruta Todeasa^10^, Cole Jensen^2^, Daniel Kim^1^, David McDonald^10^, Denise Shepard^10^, Edward Courchaine^11^, Elizabeth B. White^2^, Eric Song^1^, Erin Silva^10^, Eriko Kudo^1^, Giuseppe DeIuliis^10^, Harold Rahming^10^, Hong-Jai Park^10^, Irene Matos^10^, Isabel Ott^2^, Jessica Nouws^10^, Jordan Valdez^10^, Joseph Fauver^2^, Joseph Lim^12^, Kadi-Ann Rose^10^, Kelly Anastasio^13^, Kristina Brower^2^, Laura Glick^10^, Lokesh Sharma^10^, Lorenzo Sewanan^10^, Lynda Knaggs^10^, Maksym Minasyan^10^, Maria Batsu^10^, Mary Petrone^2^, Maxine Kuang^2^, Maura Nakahata^10^, Melissa Campbell^6^, Melissa Linehan^1^, Michael H. Askenase^14^, Michael Simonov^10^, Mikhail Smolgovsky^10^, Nicole Sonnert^1^, Nida Naushad^10^, Pavithra Vijayakumar^10^, Rick Martinello^3^, Rupak Datta^3^, Ryan Handoko^10^, Santos Bermejo^10^, Sarah Prophet^15^, Sean Bickerton^11^, Sofia Velazquez^14^, Tara Alpert^13^, Tyler Rice^1^, William Khoury-Hanold^1^, Xiaohua Peng^10^, Yexin Yang^1^, Yiyun Cao^1^, Yvette Strong^10^

^10^Yale School of Medicine, New Haven, CT, USA.

^11^Department of Biochemistry and of Molecular Biology, Yale University School of Medicine, New Haven, CT, USA.

^12^Yale Viral Hepatitis Program; Yale University School of Medicine, New Haven, CT, USA.

^13^Yale Center for Clinical Investigation, Yale University School of Medicine, New Haven, CT, USA.

^14^Department of Neurology, Yale University School of Medicine, New Haven, CT, USA.

^15^Department of Molecular, Cellular and Developmental Biology, Yale University School of Medicine, New Haven, CT, USA.

**Extended Data Fig**.**1** | **Correlation analysis of virus-specific antibodies and age, sex and BMI. a-e**, Plasma reactivity to S protein and RBD by ELISA. **a**, Anti-S IgM and IgG of total COVID-19 patients regardless of disease severity. Patients, IgM (n = 143); IgG (n = 143). **b**, Anti-RBD IgM and IgG of total COVID-19 patients regardless of disease severity. Patients, IgM (n = 99); IgG (n = 100). Each color dot represents a single individual at its maximum antibody titer over the disease course. Dashed line indicates HCW average values (limit threshold). HCW, Anti-S IgM (n = 21); IgG (n = 87); HCW, Anti-RBD IgM (n = 21); IgG (n = 21). IgG levels of Anti-S (left) or Anti-RBD (right) by **(c)** age, **(d)** sex and **(e)** BMI. Each dot represents a single individual at its maximum antibody titer over the disease course. Horizontal bars indicate mean values. OD, optical density at 450 nm (OD_450 nm_). F, females; M, males. **f**, IgG levels of Anti-S (left) or Anti-RBD by age and sex. Longitudinal analysis over time. Regression lines are shown with shading representing 95% CI.

**Extended Data Fig**.**2** | **SARS-CoV-2 viral load and disease severity. a**, Left, Viral load measured by nasopharyngeal swabs plotted as log10 of genome equivalents in non-hospitalized and hospitalized, moderate and severe COVID-19 patients. (N-hospitalized, n=10; moderate, n = 97; severe, n = 65). Each dot represents a single individual at its maximum viral titer over the disease course. Dashed line indicates threshold for positivity. Right, Average of days from symptom onset (DfSO) comparison between groups. N-hospitalized, non-hospitalized.

**Extended Data Fig**.**3** | **Overview of cellular immune profiles in COVID-19 patients. a-b**, Immune cell subsets of interest, plotted as a percentage of a parent population as **(a)** aggregate and **(b)** continuously over time according to the days of symptom onset for discharged or deceased patients. **c**, Immune cell subsets comparison in discharged or deceased patients. Negative controls (HCWs) are shown in black. Each dot represents a single individual at its maximum antibody titer over the disease course. Grey bars indicate mean values. **c**, Longitudinal data plotted over time continuously. Regression lines are shown as light blue (discharged), purple (deceased) and red (High neutralizers). Shading represents 95% CI and are coloured accordingly. (HCW, n=49; Discharged, n=122; Deceased, n=15). CD4Tfh, follicular helper T cells. ASC, antibody secreting cells. US, unswitched. CS, class switched.

**Extended Data Fig**.**4** |. **Virus-specific antibodies and viral load correlation with PRNT50. a-b**, Neutralization capacity among **(a)** total COVID-19 patients with high anti-S IgG levels (O.D.>1,4) or between mild (orange), moderate (purple) and severe (pink) at the experimental sixfold serially dilutions (from 1:3 to 1:2430). Total patients, n=63; Moderate, n=45; Severe, n=19. **c**, Levels of IgG (left) Anti-S, (middle) RBD and (right) viral load between high neutralizers, deceased and discharged patients. The indicated levels were measured at the average day from symptom onset in which each group reach 50% of neutralization at each experimental serum dilution as specified in Figure 3f. HN, high neutralizers.

**Extended Data Fig. 5** | **Gating strategies**. Gating strategies are shown for the key cell populations described in Figure 1f and Extended Data Figure 3. **a**, Leukocyte gating strategy to identify lymphocytes and granulocytes. **b**, T cell surface staining gating strategy to identify CD4 and CD8 T cells, TCR-activated T cells, follicular T cells, and additional subsets. **c**, B cell surface staining gating strategy to identify B cells subsets.

**Extended Data Tables 1-2** | **Cohort demographics using two different stratification criteria. a**, IMPACT patients and Connecticut National Guard members (mild only) stratified by disease severity. Exact counts are displayed in each cell, with standard deviations alongside. Percentages of total, where applicable, are provided in parenthesis. In cases were specific demographic information was missing, the total number of patients with complete information used for calculations is provided within the cell. **b**, IMPACT patients stratified according to their assignment as high neutralizers, discharged patients, or deceased patients. As before, exact counts are displayed in each cell, with standard deviations alongside. Percentages of total, where applicable, are provided in parenthesis. In cases were specific demographic information was missing, the total number of patients with complete information used for calculations is provided within the cell.

## Links

Lilly statement regarding NIH’s ACTIV-3 clinical trial. Lilly https://www.lilly.com/news/stories/statement-activ3-clinical-trial-nih-covid19 (26 October 2020).

May, B. Regeneron halts enrollment of critically ill patients in a COVID-19 antibody trial. BioSpace https://www.biospace.com/article/regeneron-halts-enrollment-in-covid-19-trial-following-safety-signal-in-critically-ill-patients (30 October 2020).

FDA authorizes monoclonal antibodies for treatment of COVID-19. United States Food and Drug Administration https://www.fda.gov/news-events/press-announcements/coronavirus-covid-19-update-fda-authorizes-monoclonal-antibodies-treatment-covid-19 (2020)

## Notes

### Competing Interest Statement

A.I. received sponsored research funding from Spring Discovery, and served as an expert panel in Adaptive Biotechnologies discussion.

### Funding Statement

This work was supported by the Womens Health Research at Yale Pilot Project Program (A.I.) [Fast Grant from Emergent Ventures at the Mercatus Center, Mathers Foundation, and the Ludwig Family Foundation, the Department of Internal Medicine at the Yale School of Medicine, Yale School of Public Health and the Beatrice Kleinberg Neuwirth Fund. IMPACT received support from the Yale COVID-19 Research Resource Fund. A.I. is an Investigator of the Howard Hughes Medical Institute. C.L. is a Pew Latin American Fellow. P.Y is supported by Gruber Foundation and the NSF. B.I. is supported by NIAID 2T32AI007517-16. C.B.F.V. is supported by NWO Rubicon 019.181EN.004.

### Author Declarations

Ethics statement This study was approved by Yale Human Research Protection Program Institutional Review Boards (FWA00002571, protocol ID 2000027690). Informed consent was obtained from all enrolled patients and healthcare workers.

