## Supplementary figures and images for "Kinetics of antibody responses dictate COVID-19 outcome"

### Supplemental figures

**Ext. Data Figure1:**

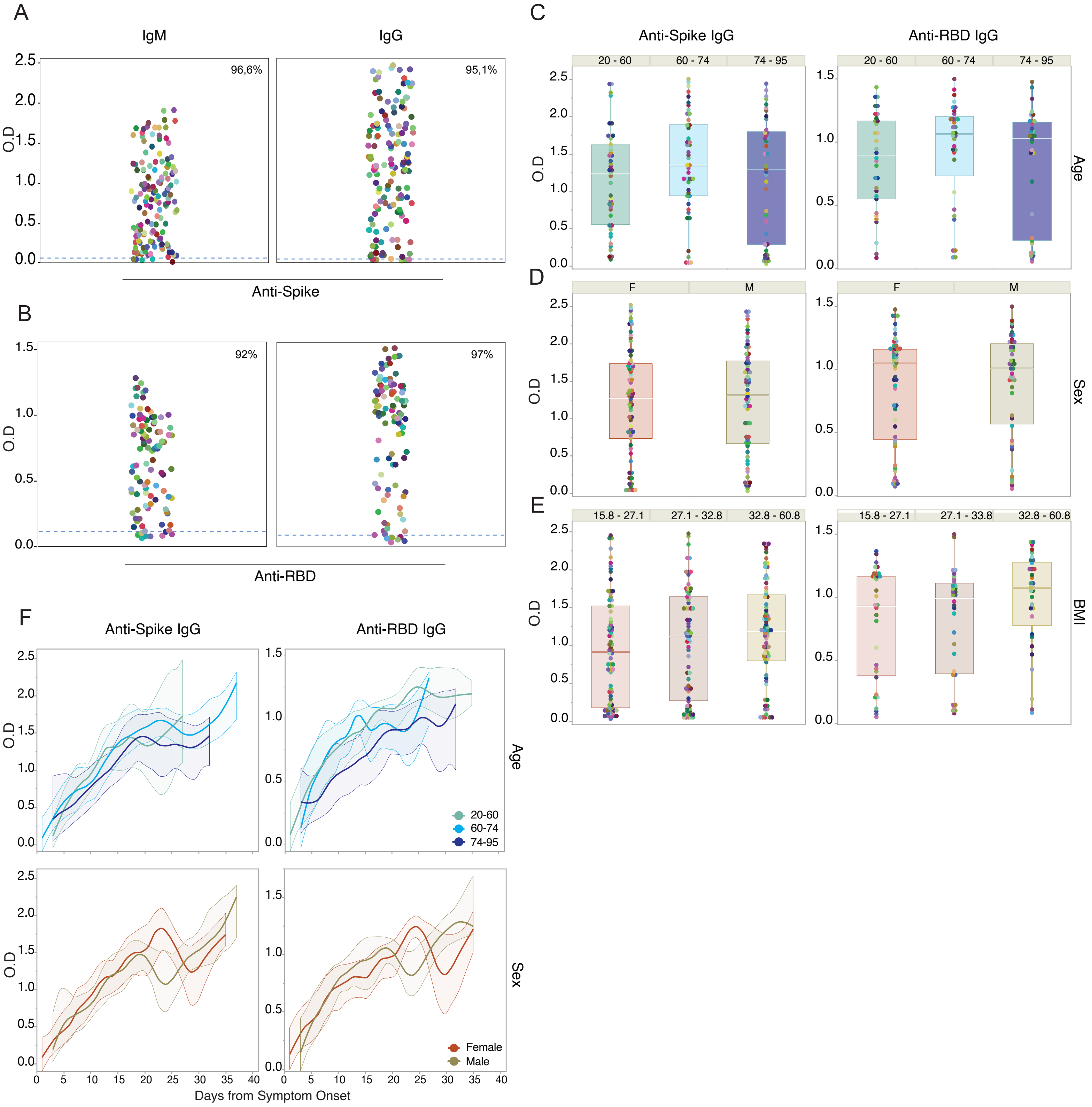

#### Ext. Data Figure2:

A

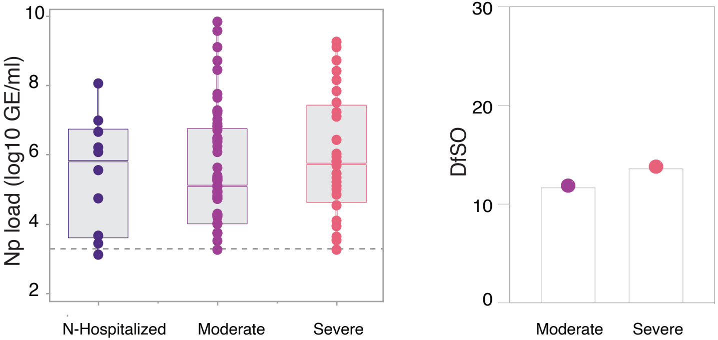

##### Ext. Data Figure3:

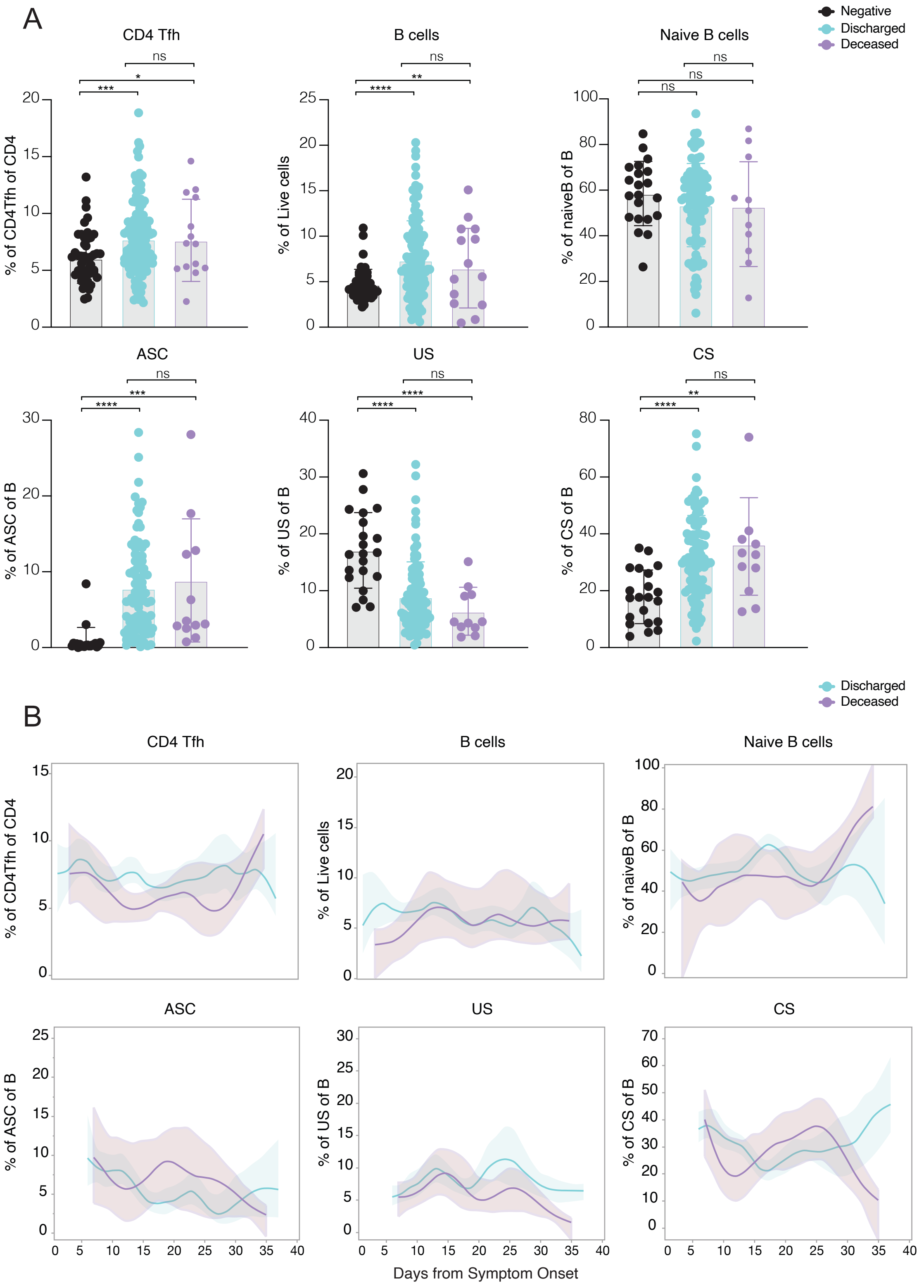

**Ext. Data Fig.4 :**

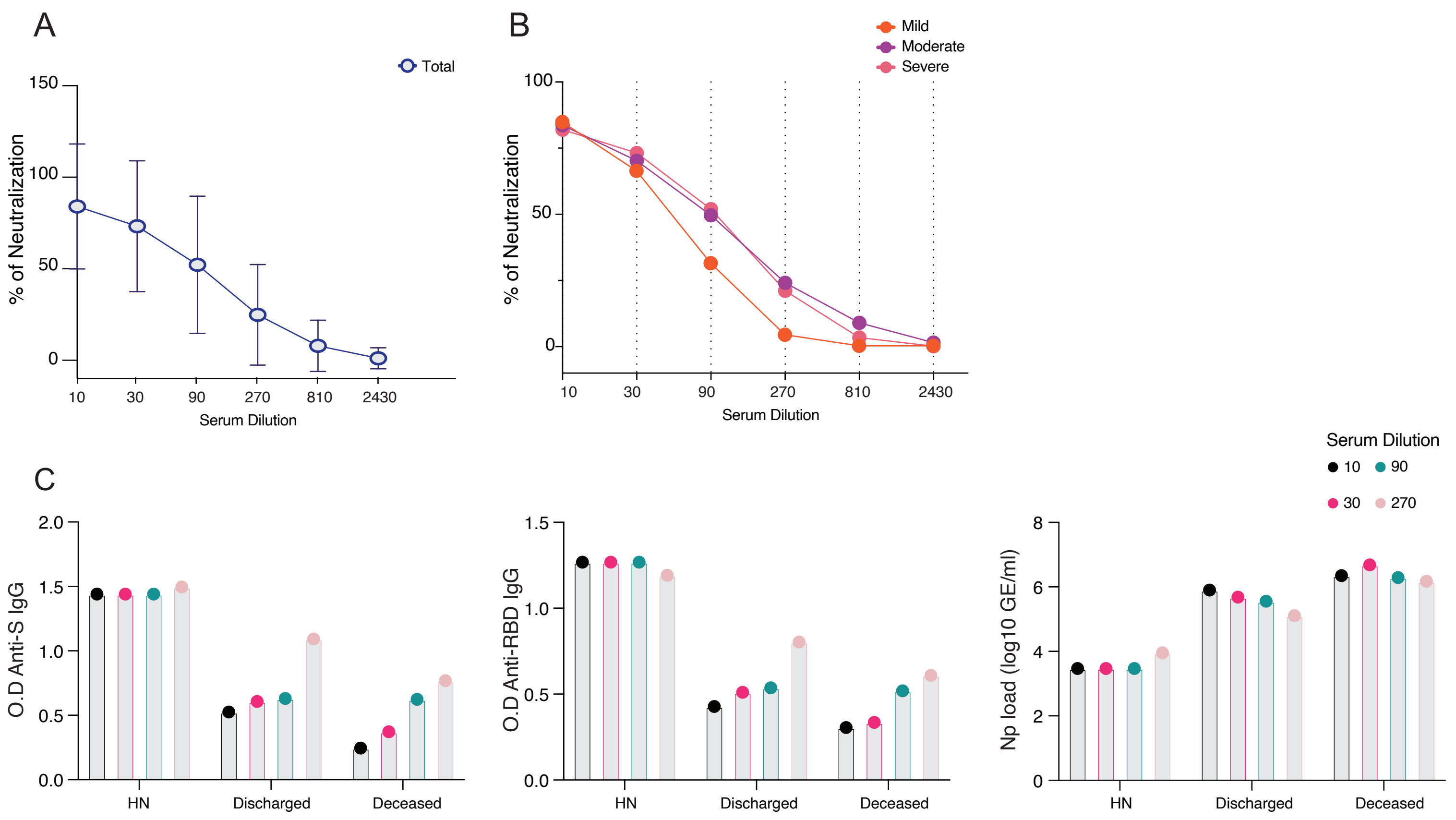

**Ext. Data Fig.5 :**

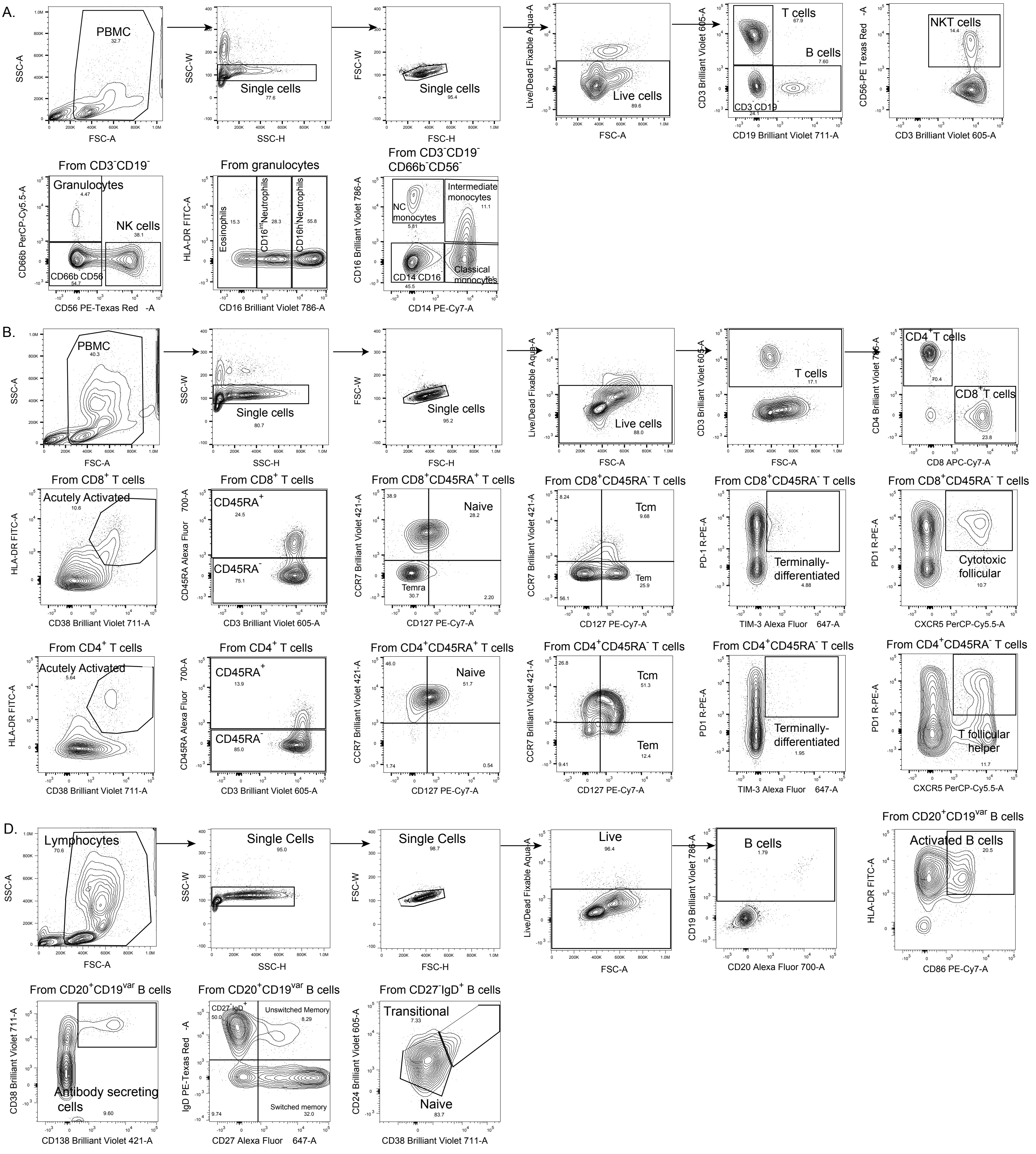
