## Supplemental Tables for "Kinetics of antibody responses dictate COVID-19 outcome"

### Extended Data Table 1 - IMPACT Cohort Total

| | Negative | Asymptomatic | Mild | Moderate | Severe | Total |
|---|---|---|---|---|---|---|
| **n** | 108 | 14 | 16 | 124 | 55 | 317 |
| **Age (years)** | 37.92 ± 11.97 | 40.57 ± 10.83 | 27.81 ± 9.85 | 62.03 ± 17.62 | 63.15 ± 17.79 | 51.33 ± 19.92 |
| **Sex (M\|F)** | 27 (25%) \| 81 (75%); n=108 | 2 (14%) \| 12 (86%); n=14 | 14 (88%) \| 2 (12%); n=16 | 58 (47%) \| 66 (53%); n=124 | 30 (55%) \| 25 (45%); n=55 | 131 (49%) \| 186 (51%); n=180 |
| **BMI** | 26.98 ± 5.95; n=114 | -- | -- | 30.2 ± 8.21; n=114 | 32.25 ± 8.8; n=50 | 29.3 ± 7.76; n=164 |

### Extended Data Table 2 - Hospitalized IMPACT Patients

| | High Neutralizers | Deceased | Discharged | Total |
|---|---|---|---|---|
| **n** | 14 | 18 | 150 | 182 |
| **Age (years)** | 56.29 ± 12.3 | 74.06 ± 19.46 | 61.75 ± 17.49 | 62.56 ± 17.75 |
| **Sex (M\|F)** | 8 (57%) \| 6 (43%); n=14 | 10 (56%) \| 8 (44%); n=18 | 70 (47%) \| 78 (53%); n=148 | 88 (49%) \| 92 (51%); n=180 |
| **BMI** | 33.89 ± 5.66; n=13 | 30.37 ± 10.57; n=18 | 30.58 ± 8.36; n=137 | 30.82 ± 8.42; n=168 |
| **COVID Risk Factors** | | | | |
| **None** | 5 (36%) | 3 (17%) | 33 (22%) | 41 (23%); n=182 |
| **Cancer (<1 year)** | 0 (0%) | 2 (11%) | 12 (8%) | 14 (8%); n=182 |
| **Chronic Heart Disease** | 3 (21%) | 5 (28%) | 41 (27%) | 49 (27%); n=182 |
| **Hypertension** | 6 (43%) | 10 (56%) | 75 (50%) | 91 (50%); n=182 |
| **Chronic Lung Disese** | 3 (21%) | 3 (17%) | 37 (25%) | 43 (24%); n=182 |
| **Immunosuppresion** | 0 (0%) | 1 (6%) | 14 (9%) | 15 (8%); n=182 |

### Extended Data Table 3 - Mild COVID-19 Cohort

| Age | n | PCR+ | Spike-ECD (OD450) | RBD (OD450) | Hospitalized |
|---|---|---|---|---|---|
| 20-29 | 13 | 7/13 (54%) | 0.90 (0.77 - 1.26) | 1.11 (0.08-2.42) | 0/13 |
| >30 | 3 | 1/3 (33%) | 1.00 (0.08 - 2.42) | 1.01 (0.54 - 1.81) | 0/3 |
